# Tailored text messaging to encourage health-protective behaviour during extreme heat in older Australians - A prototype and feasibility randomised controlled trial

**DOI:** 10.64898/2026.08.02.26359524

**Authors:** Hania Rahimi-Ardabili, Kalissa Brooke-Cowden, Anastasia Chan, Stephen Parnis, Olivia Bell, Lai Heng Foong, Enrico Coiera

## Abstract

**Introduction:** Extreme heat increasingly threatens older adults, particularly those with chronic conditions, yet generic heat-health advice may not be sufficiently timely or relevant to individual needs. This feasibility study describes a prototype and assesses the feasibility of a location-triggered, disease-specific heatwave short message service (SMS) intervention tailored to common heat-vulnerability conditions, compared with generic heatwave SMS advice.

**Methods:** Mixed-methods feasibility study comprising a parallel two-arm 1:1 randomised controlled trial and post-heatwave focus groups. Community-dwelling Australians aged ≥65 years in New South Wales, Victoria or South Australia with at least one eligible chronic condition (cardiovascular diseases, respiratory conditions, diabetes, and chronic kidney diseases) and a smartphone were recruited in summer 2026. Based on an initial codesign, participants received a “prepare” SMS after enrolment and, when Bureau of Meteorology heatwave warnings were triggered, messages before, during and after heatwaves. Control participants received generic “standard care” heat-health advice; intervention participants received condition-tailored messages and could request additional information via SMS codes. Outcomes were collected via baseline and post-heatwave surveys and thematic analysis of focus groups.

**Results:** Seventy-three participants enrolled (36 control; 37 intervention); attrition was 9.6%. Intervention engagement was strong: 61% requested additional information, with frequent free-text replies and multi-condition requests indicating preference for more conversational interaction. Eight participants were heatwave-exposed and completed post-heatwave surveys (4 per arm), with a high usability score (median of 85/100). Among these 8 participants, 7 reported adopting heat-protective health behaviours; the most common were drinking more water (6/7). More total actions were reported in the intervention group (11 vs 8). No adverse effects were reported.

**Conclusion:** A location-triggered, disease-tailored heatwave SMS system for older adults with chronic conditions was feasible, acceptable and highly usable, with high engagement and no harms. Findings support a larger trial and suggest benefits from tailored messaging.

## Introduction

Extreme heat events (heatwaves) attributable to climate change are occurring with greater frequency and intensity, posing a serious health threat (1). Older adults are at particular risk. Heat can further exacerbate underlying chronic conditions such as cardiovascular and respiratory conditions (2), leading to older adults being “doubly vulnerable.” This risk is compounded by low heat risk perception amongst older adults, which can reduce uptake of heat protective behaviours (3). Underestimating the threat of extreme heat by older adults can create a risk gap, leading to their dismissal of heat health warnings and a greater likelihood of adverse health impacts (Abrahamson, 2009; Erens, 2021).

(4, 5). Age is considered the most critical risk factor for morbidity and mortality associated with extreme heat exposure. Watts et al., (2020) report an additional 475 million heat exposure incidents for vulnerable populations in 2019 - a sharp increase from the record-breaking 160 million in 2016 (6). With rapid growth in exposure risk, heat-related deaths in older adults have consequently risen by around 85% over the past several decades (5, 7) as evidenced in major heatwaves in Europe, Russia and Asia (8). In parallel with increasing heatwave events, a global demographic shift towards ageing populations is also occurring, characterised by higher proportions of adults aged over 65 years (7).

Adverse health effects arising from heat exposure, however, are preventable. Heat-health warning systems have been developed internationally to mitigate avoidable harms by improving awareness, preparedness and prompting protective actions (9, 10). The extent to which individuals take appropriate protective action depends on their perceived risk and their confidence in knowing their needs during extreme heat events. In their 2021 Heat Health Action Plan, the World Health Organisation (WHO) reviewed heat action plans and health system responses, suggesting improvements to heat warning systems through the use of individual-level data and better heat risk communication targeted towards at-risk, vulnerable populations (5). Communication strategies are therefore a critical component of heat health action plans (HHAPs). Currently, most HHAPs contain generic, population-based messaging, which may lack efficacy if it is not timely, accessible, consistent, trusted, or sufficiently relevant to the recipient’s circumstances (5).

Tailored text messages can provide targeted actionable advice that is timely, giving older adults adequate notice to prepare for extreme heat. Tailored text messaging interventions have effectively improved a range of health behaviours including smoking cessation (11), medication adherence and lifestyle modifications (12), but have not been applied widely to heat risk communication. As the population ages and heat exposure rises, the associated health burden is expected to grow, increasing the urgency for effective and equitable prevention strategies. Effective community heat management requires understanding barriers to heat-risk information reaching vulnerable groups (13).

This study evaluates the feasibility of deploying a prototype disease- and location-specific heatwave short message service (SMS) intervention, compared with generic heatwave SMS alerts, for older Australians (≥65 years). The feasibility trial evaluates methodological feasibility, early engagement, and participant acceptability and trial experience. With mobile phone ownership amongst older Australian adults around 70% (14), personalised heat risk communication delivered via text-based messaging may offer an effective and scalable approach.

## Methods

### Trial design

A mixed-methods approach was selected for this feasibility study, including a parallel double-arm 1:1 randomised controlled trial (RCT) followed by a focus group study. Participants in the intervention arm received tailored disease-specific heatwave text messages, and the control arm received “standard care” general heatwave text. This trial was registered with the ANZCTR (ACTRN12625000795493) on 25/07/2025 and has been conducted in accordance with the CONSORT 2025 guidelines.

### Co-design-informed evaluation

We conducted a brief hybrid co-design-informed evaluation session of various SMS message options. A small group of volunteers (n=6) attended a single 1-hour session in which they received a series of SMS message options and provided immediate feedback. Each SMS series differed in presentation features (for example: readability, message length, frequency). Participants indicated which option they preferred and reported their experience of the messages. Structured items captured perceptions of readability/clarity, appropriate length, frequency, and language. Brief open-ended prompts captured reasons for preferences and suggestions for improvement. Responses were recorded and used to refine message design and characteristics. Based on the feedback, message content was split into multiple shorter messages sent at different phases of the heatwave (i.e. preparation phase, before, during and after a heatwave), with an optional ‘read more’ feature for additional information.

### Participant recruitment

Participants were recruited during the Australian summer in early January 2026, and the tailored text messaging intervention was in operation from 25 Jan 2026 to 28 Mar 2026 across three Australian states: New South Wales (NSW), Victoria and South Australia (SA). A multi-channel recruitment strategy used advertising via an online research recruitment registry (JoinUs) and email invitations to individuals who had previously expressed interest in participating in this research. Eligibility included older Australian smartphone owners who had at least one medical condition (Table 1). A random subgroup of RCT participants from diverse backgrounds was invited to participate in semi-structured focus groups.

**Table 1.**
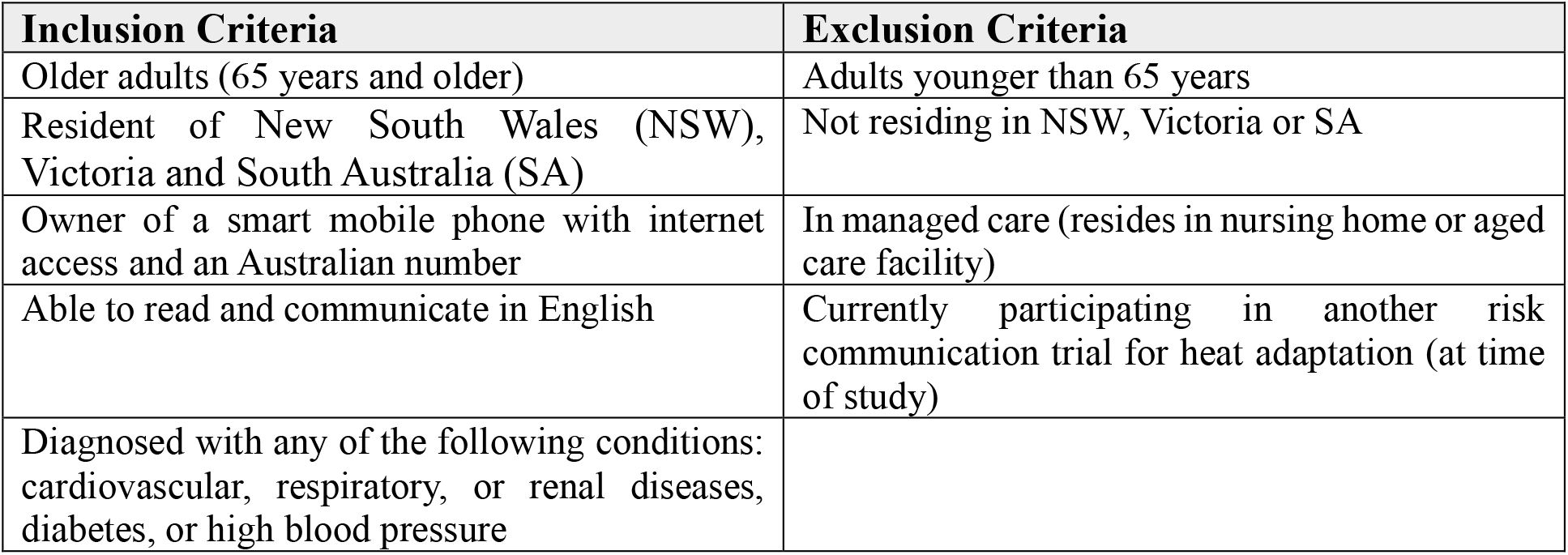
Inclusion Criteria.

### Intervention

The intervention was triggered when a heatwave event was predicted for a region within the study States, using the Australian Bureau of Meteorology (BoM) heatwave definition and its Heatwave Warning Service. This aligns with the WHO’s recommended risk communication occurring 1-3 days before a heat event. Once triggered, the SMS system sent four sets of heat-health messages to participants in the target region, varying according to their group allocation. Messages were delivered in four sets: a “prepare” message (after enrolment, but before any heatwave), and then messages sent before (up to one day prior to the heatwave), during, and after a predicted heatwave. The participants in the control arm received “standard care,” which was a general set of health actions for heatwaves (e.g., how to stay cool). Participants in the intervention arm received messages tailored to four specific medical conditions of 1) cardiovascular diseases, including high blood pressure, 2) asthma or chronic obstructive pulmonary disease (COPD), 3) chronic kidney diseases or 4) Diabetes. Participants in the intervention group could elect to receive a follow-up text message for additional information on disease-specific information or general cooling strategies and preparedness by responding with defined codes listed in the text, allowing for further customised content.

### Message library development

Message development occurred in consultation with three emergency medicine clinicians, including two senior emergency physicians (SP, LHF) and one emergency medicine registrar (OL), to identify priority heat-vulnerability diseases for the purpose of the feasibility study. Informed by peer-reviewed articles, clinical guidelines and action plans such as Heat Health Action Plans, we developed evidence-based recommendations and strategies tailored to cardiovascular diseases, including hypertension, asthma, COPD, chronic kidney diseases and diabetes during heatwaves.

### Message framing

We reviewed multiple risk communication guidelines to inform message framing, including the WHO guidelines for communicating risk in health emergencies (15) and Australian Government Department of Health Emergency Warning guidelines (16) and developed a framework (Supplementary file Appendix 1) emphasising accessibility, actionability, and trustworthiness. We reviewed multiple risk communication guidelines to inform message framing, including the WHO guidelines for communicating risk in health emergencies (15) and Australian Government Department of Health Emergency Warning guidelines (16) and developed a framework (Supplementary file Appendix 1) emphasising accessibility, actionability, and trustworthiness. We reviewed multiple risk communication guidelines to inform message framing, including the WHO guidelines for communicating risk in health emergencies (15) and Australian Government Department of Health Emergency Warning guidelines (16) and developed a framework (Supplementary file Appendix 1) emphasising accessibility, actionability, and trustworthiness. Accordingly, the selected message format was: [Trial NAME]: [TOPIC]. [ACTION]. [RISK/SUPPORTING INFO]. [RESOURCES]. [CLOSING]. Figure 3 is an example of tailored text messaging offering guidance on respiratory conditions with recommended actions during extreme heat, drawn from the literature (17, 18). A linguistics/conversation analyst (Bedside Manners) was consulted to refine message clarity and brevity.

**Figure 1.**
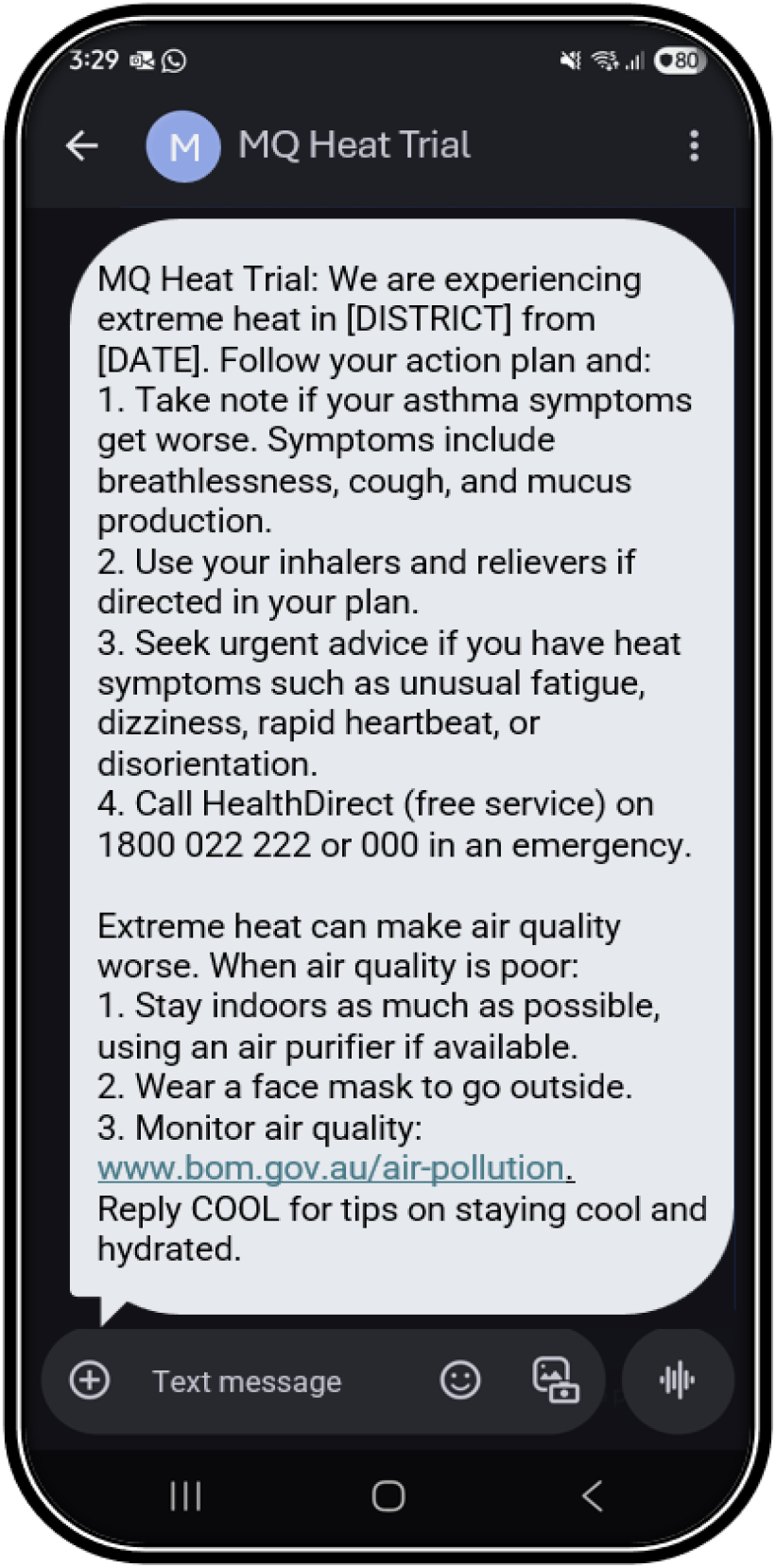
Screenshot example of respiratory advice during the heatwave

### SMS engine development

We employed commercial off-the-shelf (COTS) technology platforms through Amazon Web Service ((19)) to:

- Aggregate and then ingest participant and weather data into secure databases. Participant data collected at recruitment included mobile number, residential postcode, and heat-related risk factors such as pre-existing health conditions and medications. Postcode mapping linked participants’ locations to weather data, enabling targeted, context-specific interventions. BoM forecast data was monitored via an anonymous FTP (File Transfer Protocol) portal and updated in real-time. The Amazon platform incorporates a middleware integration layer that translates BoM heatwave warning data into intervention triggers. Automated monitoring and redundancy safeguards support real-time data integrity and operational continuity in the event of any changes.
- Tailor messages: A rule engine used participant and heat event data to select and tailor SMS messages from a message library. The rule engine used participant and weather data to select between different message options specific to health conditions and comorbidities. A text engine then constructed the tailored text message using an evidence-based message library (see above). Finally, a message scheduler decided when to send messages based on data such as a participant’s location.
- Distribute: Once scheduled, tailored SMS texts were transferred securely to an Australian-located Amazon text-messaging aggregator for distribution to trial participants (see Figure 2).

**Figure 2.**
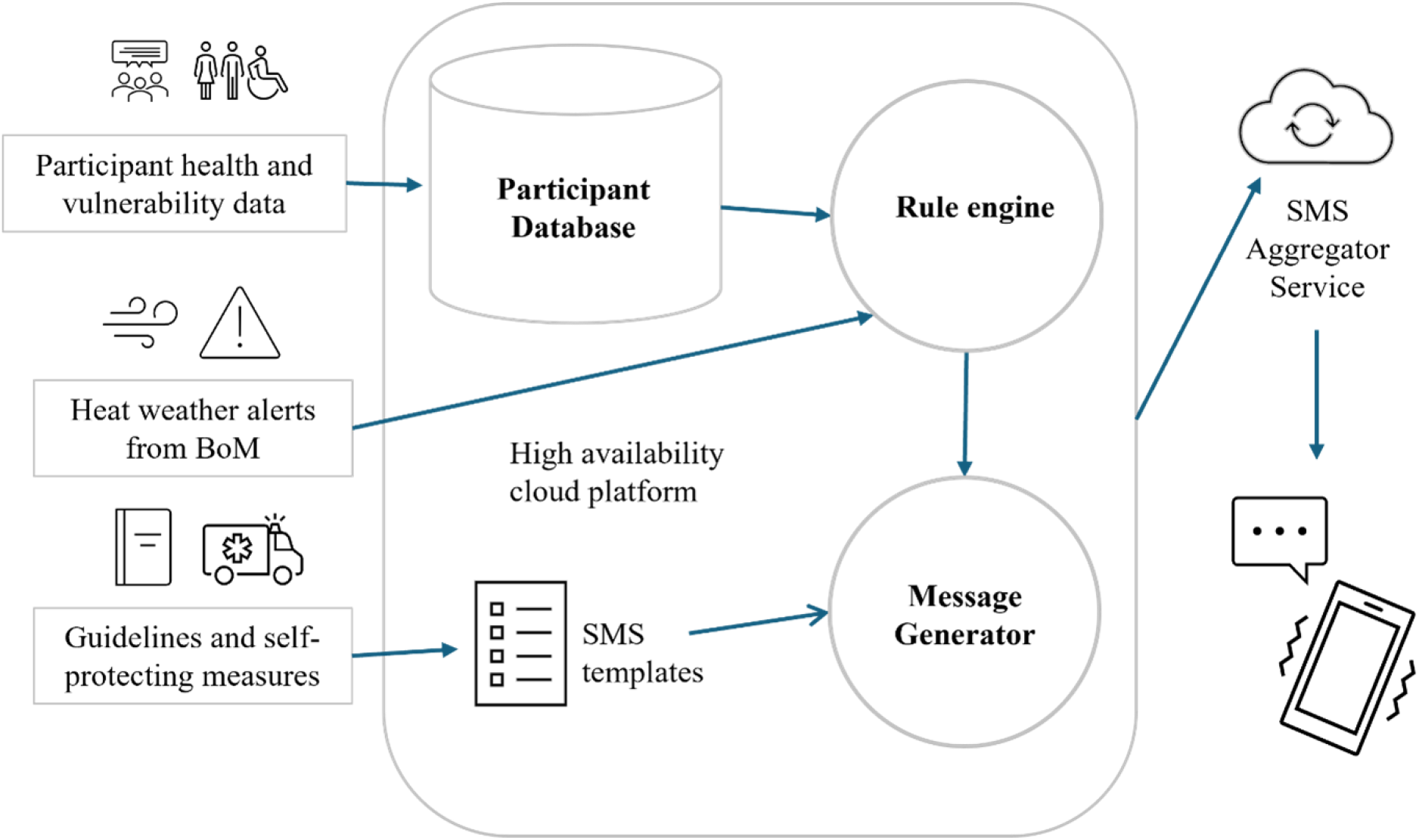
The COTS platform for tailored SMS broadcasting.

### Measures

Participant information was collected using self-reported data via online surveys (REDCap) at baseline and post-heatwave follow-up (see Supplementary file 2 for questions and sources). Outcomes included heat-related symptoms (e.g., dizziness), health service utilisation (e.g., hospital or general practitioner visits) at baseline and post-heatwave follow-up, adherence to heat-related directives, adverse effects, and intervention acceptability at post-heatwave follow-up. A heat symptoms survey was adapted from prior questionnaires used in studies such as Mehiriz et al. 2018 (see Supp File 2). Usability was assessed via the System Usability Score (SUS) adapted for this study. Interaction with the SMS system was assessed using participants’ replies, either as free text or in response to offers of additional information. Online focus group data collected post-heatwave were used to assess overall experience, feasibility, and acceptability, and also harms, barriers and facilitators. Focus group sessions were conducted via Microsoft (MS) Teams, using the recording and transcription functions. Table 2 presents the timing of participant information collection and outcome measures.

**Table 2.**
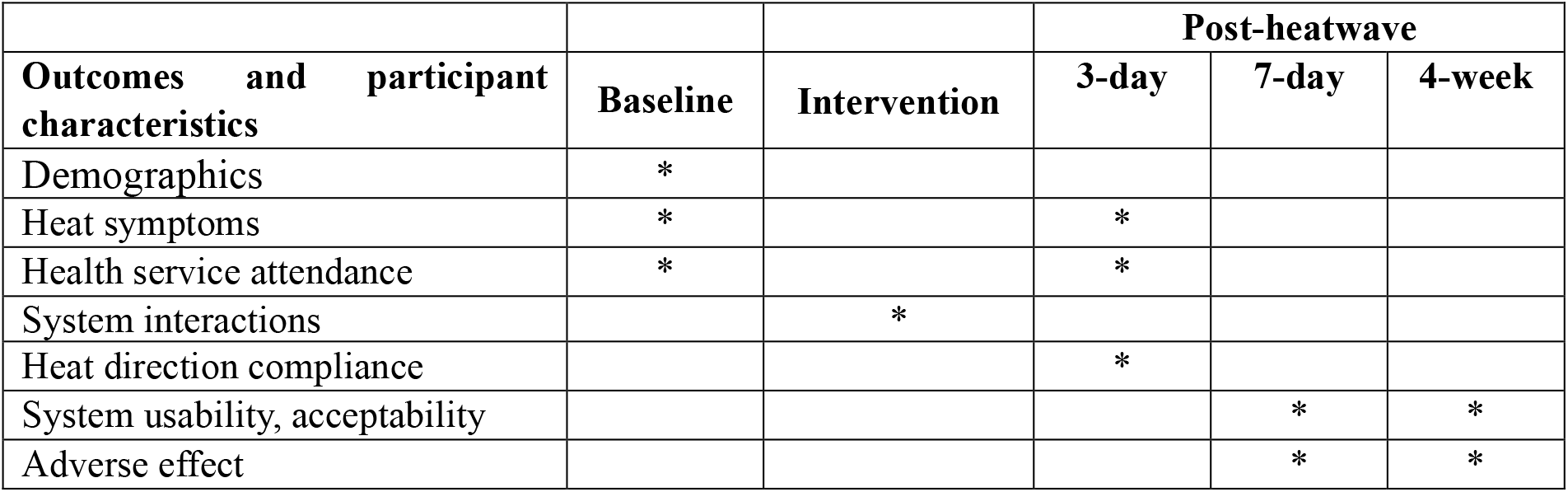
Outcomes, participant characteristics and their assessment timepoints.

### Randomisation and allocation concealment

Participants were blinded and unaware of their group allocation. A computer-based random number generator was utilised to allocate participants to either intervention or control groups. Allocation concealment was ensured using sealed envelopes prepared in advance by an independent individual not involved in the research containing the corresponding group code and opened only after a participant’s enrolment was confirmed.

### Data Analysis

*RCT:* Baseline characteristics and demographic factors (age, sex, education level, location and pre-existing health conditions) were summarised for all participants. Results are presented as counts (percentages) for categorical variables and as medians (interquartile ranges, IQRs) for continuous variables. Baseline group-comparison test methods for assessing balance are provided in Supplementary File A2.

System usability using the System Usability Scale (SUS) was calculated by summing item scores (odd items scored as “response−1”; even items scored as “5−response”) and multiplying the total by 2.5 to yield a score from 0 to 100. Given the small number of participants who experienced a heatwave and provided post-heatwave data, between-group comparisons of usability and symptom-related outcomes were treated as exploratory and are reported using descriptive summaries rather than formal hypothesis tests. All statistical analyses were conducted using IBM SPSS Statistics 31.

*Focus group*: Data from the focus groups were analysed using Braun and Clarke’s (2006) reflexive thematic analysis to identify key themes. De-identified transcripts were coded inductively line-by-line by one researcher (HRA) to generate initial codes and categories, which were iteratively reviewed and refined through team discussions to develop and achieve consensus on key themes.

## Results

### Sample and participation

Of 145 individuals expressing interest, 54 were ineligible, and 18 did not consent, resulting in 73 enrolled participants (36 control; 37 intervention), and a response rate of 80.2% (73/91). Participants had a median age of 70 years (IQR 9.44) and were recruited from New South Wales (48%), Victoria (37%), and South Australia (15%). Seven participants withdrew or were lost to follow-up (5 control; 2 intervention), resulting in an attrition rate of 9.6% (7/73). Consort diagram Figure 3. Eight participants attended one of three focus groups, with each session averaging 30 minutes, selected from various backgrounds; their demographics are provided in Table A1 (supplementary file).

### Participant characteristics

The study sample comprised a diverse cohort of older adults with varied health, socioeconomic, and demographic characteristics (Table 3). Overall, health status was moderate, with 50% (n = 37) reporting good or very good health and 30% (n = 23) reporting fair health. The cohort was predominantly female (70%, n = 51) and largely originating from Oceania (i.e. Australia and Pasifika; 67%, n = 49). Socioeconomic variation was evident, with half (50%, n = 38) reporting low to middle-low income (AUD 18,201–78,000) and half (50%, n = 37) holding a university qualification. Most participants were retired (72%, n = 53) or semi-retired (16%, n = 12), and 34% resided in rural or regional areas. A notable proportion required additional support, with 20% (n = 15) receiving carer assistance and 33% (n = 24) managing disabilities. 12% (n = 9) were from non-English speaking backgrounds.

**Table 3.** Participant characteristics and living arrangement.

|  |  | **Control**<br>**(n=36)** | **Intervention**<br>**(n=37)** | **Total** |
| --- | --- | --- | --- | --- |
| **Health status** | **Excellent** | **0** | **0** | 0 |
|  | **Very good** | 10 | 11 | 21 |
|  | **Fair** | 11 | 12 | 23 |
|  | **Good** | 8 | 8 | 16 |
|  | **Poor** | 6 | 6 | 12 |
|  | **Not sure** | 1 | 0 | 1 |
| **Gender** | **Male** | 11 | 11 | 22 |
|  | **Female** | 25 | 26 | 51 |
| **State** | **NSW** | 17 | 18 | 35 |
|  | **VIC** | 14 | 13 | 27 |
|  | **SA** | 5 | 6 | 11 |
| **Annual household**<br>**income (before tax)** | **18,200 AUD or less** | 0 | 4 | 4 |
|  | **18,201 - 45,000 AUD** | 15 | 7 | 22 |
|  | **45,001 - 78,000 AUD** | 7 | 10 | 16 |
|  | **78,001 - 103,000 AUD** | 5 | 9 | 14 |
|  | **103,001 AUD and above** | 5 | 2 | 7 |
|  | **Prefer not to say** | 4 | 6 | 10 |
| **Education level** | **High school or below** | 4 | 5 | 9 |
|  | **Certificate or Diploma** | 11 | 15 | 26 |
|  | **Bachelor's degree** | 12 | 6 | 18 |
|  | **Postgraduate degree** | 9 | 10 | 19 |
|  | **Prefer not to say** | 0 | 1 | 1 |
| **Aboriginal and/or**<br>**Torres Strait**<br>**Islander** | **Yes, Aboriginal** | 0 | 1 | 1 |
|  | **Not sure/don't know** | 1 | 0 | 1 |
|  | **No** | 35 | 36 | 71 |
| **Country/region** | **Oceania and Antarctica** | 25 | 24 | 49 |
|  | **North-West Europe** | 7 | 3 | 10 |
|  | **Southern and Eastern**<br>**Europe** | 1 | 2 | 3 |
|  | **South-East Asia** | 2 | 2 | 4 |
|  | **North-East Asia** | 0 | 1 | 1 |
|  | **Southern and Central Asia** | 1 | 1 | 2 |
|  | **Sub-Saharan Africa** | 0 | 2 | 2 |
|  | **Not sure/don't know** | 0 | 1 | 1 |
|  | **Prefer not to say** | 0 | 1 | 1 |
| **Employment and retirement** | **Retired (not working)** | 23 | 30 | 53 |
|  | **Semi-retired (part-time)** | 7 | 6 | 13 |
|  | **Working full-time** | 4 | 1 | 5 |
|  | **Other (volunteering)** | 2 | 0 | 2 |
| **Location** | **Urban or suburban** | 22 | 26 | 48 |
|  | **Regional** | 8 | 9 | 16 |
|  | **Rural or remote** | 6 | 3 | 9 |
| **Living arrangement** | **Live alone** | 10 | 12 | 22 |
|  | **Live with partner or family** | 25 | 23 | 48 |
|  | **Shared accommodation** | 1 | 2 | 3 |
| **Carer support or assistance** | **Yes** | 4 | 11 | 15 |
|  | **No** | 32 | 26 | 58 |
| **Physical or mental disabilities** | **Yes, mental** | 0 | 1 | 1 |
|  | **Yes, physical** | 11 | 12 | 23 |
|  | **No** | 25 | 24 | 49 |
| **Non-English speaking background** | **Yes** | 2 | 7 | 9 |
|  | **No** | 34 | 30 | 64 |

NSW, New South Wales, VIC, Victoria; SA, South Australia

**Fig 3.**
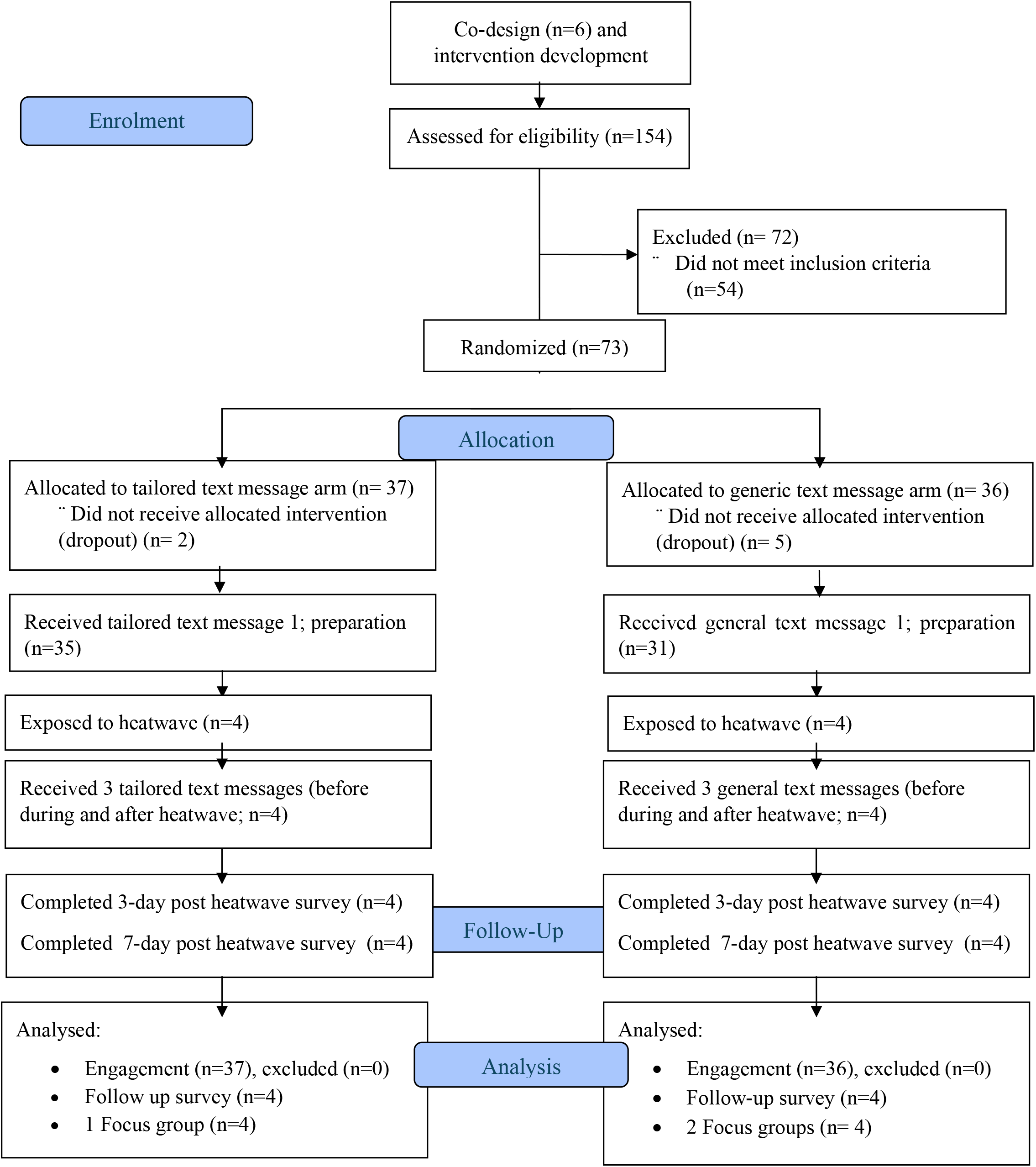
Consort flow diagram.

A range of heatwave vulnerable conditions was represented, with comorbidities common (59%, n = 43). Cardiovascular and metabolic conditions were most prevalent (81%), including high blood pressure (63%), followed by asthma (42%), diabetes (36%), COPD (8%), and chronic kidney disease (10%). Other commonly reported conditions included neurological, mental health (22%) and musculoskeletal (21%) disorders.

### Baseline group comparison

At baseline, there were no significant differences between the intervention and control groups, or between enrolled and dropout participants, across education, income, living arrangement, carer support, gender distribution, multimorbidity, overall health, employment status, disability status, home locality, or symptoms reported (ps ≥ 0.07). See Table A2 in the supplementary file for baseline comparison.

### Interaction (intervention group only)

Participants’ interaction frequency suggested good uptake of the SMS system during the trial. Intervention participants interacted with the SMS system 72 times, and 61% of participants (n= 22) requested additional information. The median additional interactions was 2 SMS requests per participant (IQR=3; range 0-10) during the preparation phase (the first set of recommendations). No interaction options were available for the control group. Out of 72 messages received, 54 were replies to an offer of preset options, and 18 were free text. More than one-third of the responses (26/72) were for condition-specific content (Box 1).

**Box 1.**
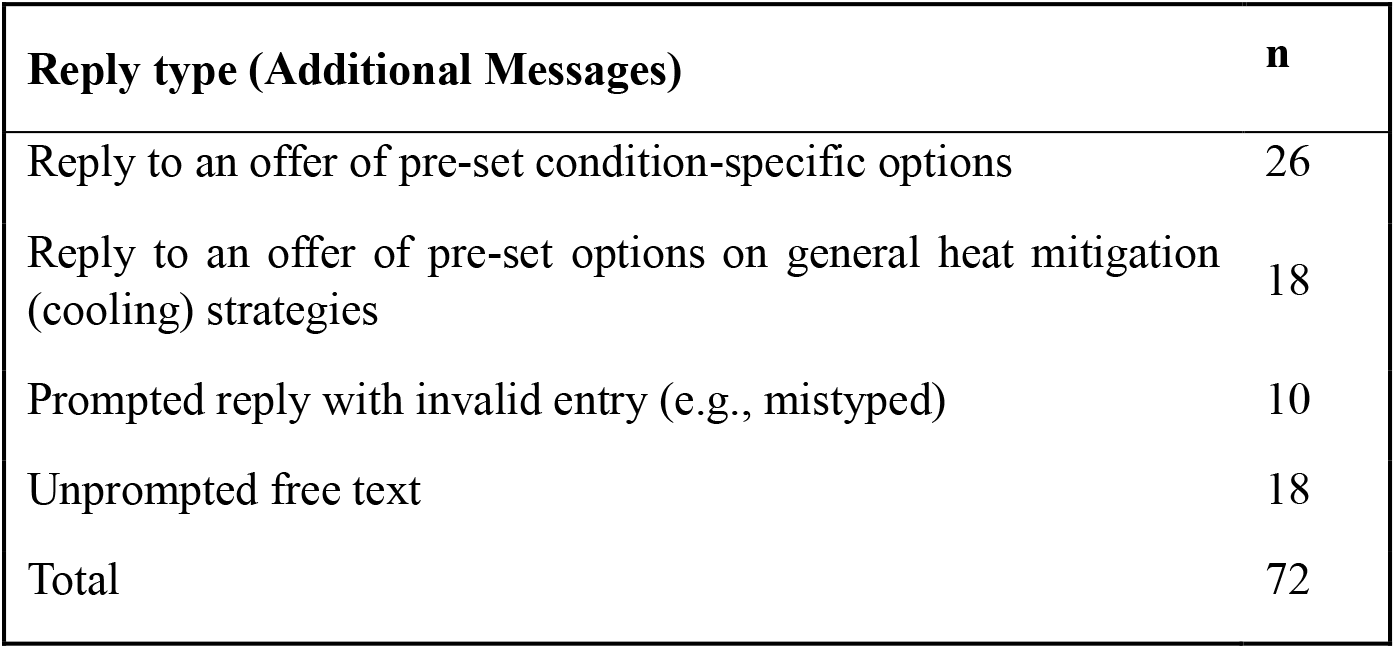
Type of information requested during the intervention (intervention group only)

All four participants who were exposed to heatwaves in the intervention group, replied to offers of additional preset options, triggering a median of three extra SMS responses (IQR=9.5; range 1-18) for a single heatwave episode. This demonstrates willingness to engage and evidence that the information being provided was perceived as valuable. Some responses suggested appreciation of the messages and enthusiasm for further tailored guidance; for example, one participant wrote, “*Ok. Thanks. So far, all covered. Looking forward to the next episode*” (male, 73 years old).

### Usability

Usability was highly rated among both groups (System Usability Scale median 85 out of 100, IQR 8.75; Grade A/top quartile), (Group breakdown Intervention (n=4), median: 83.75, IQR: 15; Control (n=4): median 85, IQR: 7.5). Participants consistently agreed that the text-messaging intervention was easy to use, well organised, and quick to learn, while strongly disagreeing that it was complex, inconsistent, or required technical support. Figure 4 illustrates the median and IQR for SUS items. Overall, the findings suggest that the intervention demonstrates high usability, and the users specified that they “felt confident” (item no.9, see Figure 4). All participants agreed that the messages provided sufficient information to act.

**Figure 4.**
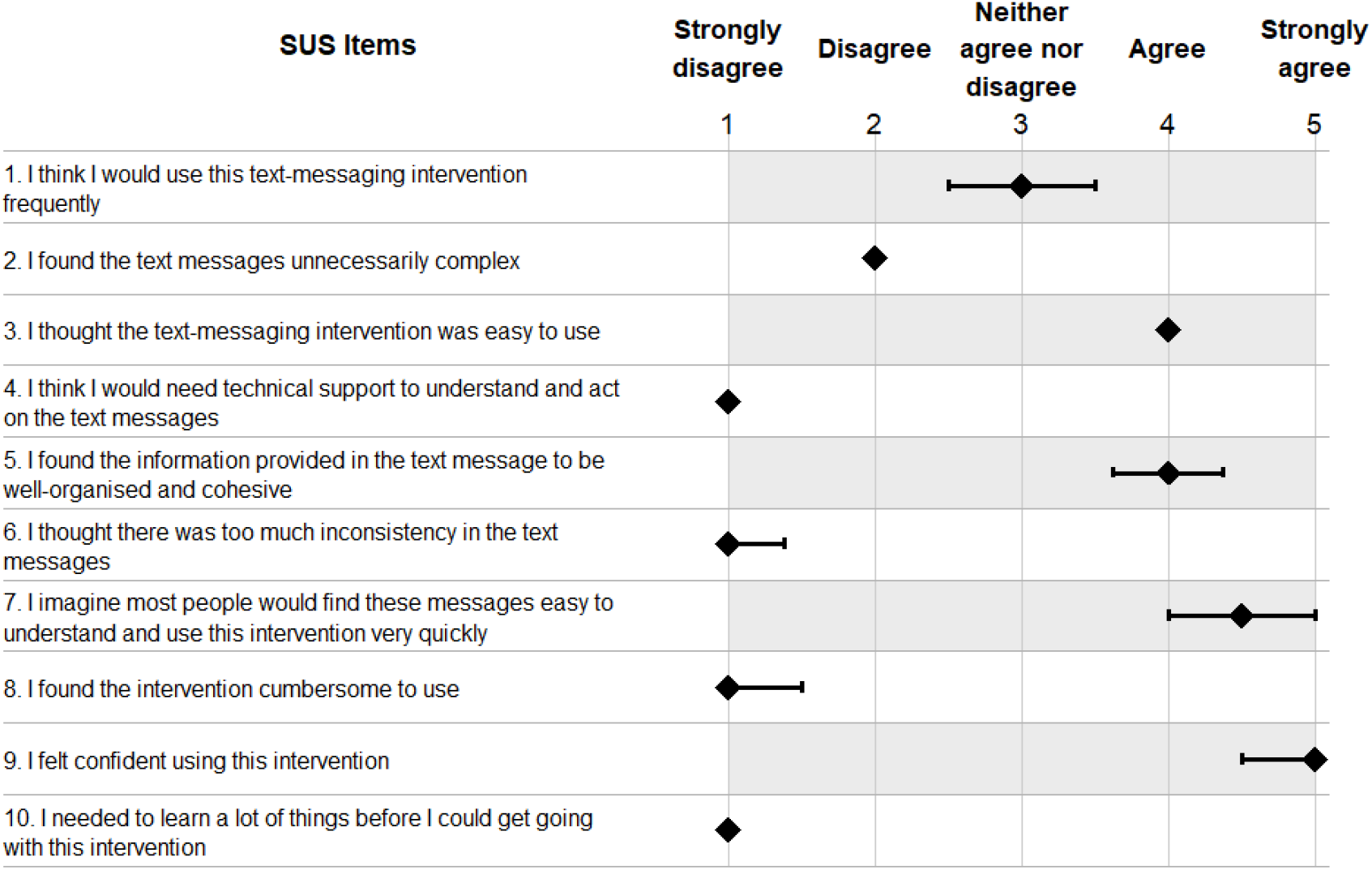
Diamonds designate the median ratings, and the lines display the interquartile range (IQR = Quartile 3 – Quartile 1). White rows indicate negatively worded items. Rating of the Heat Trial text intervention using the System Usability Scale (SUS; n=8)

### Behavioural Responses to Message

Across both groups (n=8), seven participants (3/4 in control; 4/4 in intervention) reported taking at least one action after receiving messages. Only a control participant reported no action due to being in an air-conditioned environment. Of the 7 participants from both groups who reported taking actions the most common behaviours were increased hydration (n = 6) and staying cool through measures such as using air conditioning or staying indoors (n = 6). Some participants also avoided physical exertion (n = 3) and monitored for heat-related symptoms (n = 2). Two reported disease-specific monitoring (n = 1) or other actions (n = 1), while no participants consulted a health provider, prepared medication storage, or used a buddy system. Overall, a slightly higher number of actions were reported in the intervention group compared to the control group (11 vs 8 actions).

### Reported symptoms and health services use

Sleeping difficulties (30%, n = 22), breathing difficulty (18%, n = 13), and dizziness/light-headedness (22%, n = 8) were the most common symptoms at baseline, while headache and trouble sleeping were most reported at 3-day follow-up (n = 8).

At baseline, 15% (n = 11) had visited a general practitioner, and 4% (n = 3) had seen a specialist in the previous 7 days; no ED or pharmacy use in the last 7 days was reported. Among those exposed to the heatwave (n=8), no participant reported using health services before, during, or after the heatwave in either group.

### Adverse effect responses

Across four adverse-effect items, all participants who were exposed to a heatwave (n = 8) reported no adverse outcomes associated with the text message intervention. All indicated they had not experienced any health issues related to the messages, had not felt unsafe, at risk, or in significant distress after acting on the advice (“*not at all*”), had not reported any adverse experiences to the study team, and had not experienced any injury or health problem directly linked to actions taken following receipt of the messages.

Focus groups participants also reported that they did not experience harm, confusion, distress, or adverse effects related to the intervention, stating *“not at all or definitely not”* (IDs 29, 34). Participants described feeling “*comfortable receiving”* (ID77, intervention group) the messages and did not report negative consequences from the information or recommended actions.

### Participants’ experience (focus groups)

Participants across both groups mostly described the intervention positively, with participants who only partially engaged due to a lack of heatwave events in their district still identifying potential value. In particular, those with comorbidities acknowledged their vulnerability to heat, underscoring that such interventions can be important. In the intervention group, participants reported increased awareness by saying “*It’s the awareness. It’s the big winner*” (ID77, intervention group) or “*It did make me think*” (ID34, control group). Preferences regarding tailoring varied. While one participant from the control group stated, “*If there were things that were specific to me, it would catch my attention*” (ID34, control group), others commonly characterised as an alert or reminder system or a “*nudge*”, with generic messaging more closely aligning with how participants conceptualised its purpose. “*It’s just like a memory jogger, which is always good”* (ID77, intervention group). Perceived benefits included timely prompts that supported immediate actions. Perceived redundancy was observed in one participant in the control group. “*I know what to do*” (ID29, control group) supporting the evidence that generic messaging may be of limited perceived usefulness for some population groups.

Message length was generally considered appropriate. Some participants thought the messages could have been shorter. “*Sometimes less is more*” (ID29, control group). Focus group findings aligned with SUS responses, showing perceived ease of use, clear language, and an appropriate tone. *“It seemed very appropriate… straightforward… polite*.*”* (ID14, control) or *“I didn’t have any problem with it*.*”* (ID70, intervention).

Participants commonly described the SMS messages as increasing “awareness” and acting as a “reminder” or “nudge”, suggesting a plausible mechanism of effect as a cue to action on hot days. Preferences for tailoring were mixed: some participants indicated that condition-specific personalisation would increase salience, whereas others viewed the system primarily as a generic alert, supporting a design that offers simple universal prompts alongside optional, on-demand tailored advice. Participants also highlighted equity constraints, noting that housing conditions and financial barriers (for example, limited access to air conditioning) can restrict the ability to enact recommended behaviours; this indicates that messaging-only approaches may need to be paired with structural supports and alternative delivery pathways to be effective and equitable. Finally, usability feedback pointed to clear design priorities for future iterations, including persistent access to information beyond a single SMS view and more semantically flexible interaction that accommodates multi-condition needs and conversational inputs.

Limitations included potential difficulty for individuals with limited technology literacy and the lack of ongoing visibility for SMS messages after they are read “*once you’ve read them, they’re gone*” (ID11, control group). In the intervention group, participants managing multiple conditions reported difficulty accessing tailored advice due to the response function restriction of a single condition pathway at a time, consistent with interactions with the system data indicating uncertainty in accessing customised messages. This highlighted the need for platforms that combine persistent message visibility with semantically flexible and conversational interaction. Suggestions for improvement centred on equity and inclusivity, including support from family or carers, “*having the carer there… then you’ll know that your message will get through*” (ID29, control group), alternative delivery pathways such as personal alarm systems and multilingual messaging options. Participants also highlighted the influence of housing conditions and financial constraints on heat-related behaviours “*we don’t all live in houses that have got air conditioning*” (ID70, intervention group). Broader dissemination through public alerts, printed materials, and healthcare settings was also suggested for inclusivity.

## Discussion

This prototype location-triggered, disease-specific heatwave SMS RCT for older Australians with chronic conditions was feasible, implementable, and highly usable among participants in surveys or focus groups—supporting acceptability of SMS-delivered prompts for heat-health action. No harms or adverse effects related to recommendations were reported. Usability was high in both arms (median SUS score of 85/100; “Grade A/top quartile”), and participants described the intervention as easy to use, clearly worded, and appropriately toned.

A range of heatwave-vulnerable conditions was represented, with comorbidities common (59%) and cardiovascular and metabolic conditions the most prevalent (81%). This suggests the approach may be relevant to many older adults living with multiple chronic diseases rather than a single diagnostic group. In the intervention arm, over one-third of additional replies (26/72) requested condition-specific information using pre-set options, indicating uptake of the condition-tailored component. Almost all participants who experienced heatwaves reported adopting some health-protective behaviours during the heatwave, such as increased hydration (n = 6), staying cool (n = 6), and monitoring for symptoms (n = 3).

Large future randomised controlled trials are needed to examine the effectiveness and generalisability of this feasibility study across multiple heat events. Future trials should test impacts on protective behaviours and heat symptoms; assess short- and longer-term changes in participants’ knowledge and preparedness; and assess the equity of reach and benefit across subgroups. The small sample and limited number of heatwave exposures in this trial restrict inferences about effectiveness across conditions and contexts. Scalability is supported by alignment with emerging trial initiatives that use tailored digital heat-health messaging (20). For example, Canada’s Heat Smart project is evaluating tailored heat-health text messages facilitated through primary care and integrated with public health heat-alert systems, to assess the impact on heat-adaptive behaviours. Embedding SMS prompting within existing delivery platforms (for example, primary care) provides a plausible pathway to expand reach and reduce implementation burden in larger trials (20).

Participant feedback suggests that tailoring may increase salience for some (“specific to me… catch my attention”), whilst others perceived alerts as a general “reminder”. Consistent with the literature (21, 22), some participants found the generic information irrelevant to them or something they ‘already knew’, emphasising the potential importance of a tailored approach.

Engagement data showed a strong appetite for two-way support (61% requested extra information) but also revealed challenges with single-pathway rules for people managing multiple conditions, which hindered participants from receiving the appropriate further tailored message requested. Frequent free-text and multi-code replies indicate users naturally communicated conversationally; moving from code-based branching to a conversational approach could interpret multi-condition queries and reduce user error.

## Limitations

Post-heatwave outcome data came from a small heatwave-exposed subgroup, and between-group comparisons and efficacy analysis were not possible. The main contribution is feasibility/usability and interaction assessments. Thus, a large-scale randomised trial is needed to assess intervention effectiveness. The sample broadly reflects older Australians in terms of retirement status, health status, and rural/regional representation, although it appears less representative in cultural and linguistic diversity. The sample is also skewed towards higher socioeconomic position, particularly higher education, and gender distribution shows a higher proportion of females than in the general older Australian population. Further, participants were recruited via an online platform, which may have resulted in a sample with higher digital literacy than the general older population, potentially limiting the generalisability of the usability and interaction findings.

## Conclusion

This location-triggered, disease-specific heatwave SMS intervention for older Australians with chronic conditions was feasible to implement, highly usable, and acceptable, with no harms reported. Perceived relevance varied, highlighting the importance of tailoring. Future large-scale trials are needed to assess the intervention’s effectiveness while being rigorously evaluated for safety.

## Data Availability

All data produced in the present study are available upon reasonable request to the authors, subject to Ethics approval.

